# Repeatability of RRate measurements in children during triage in two Ugandan hospitals

**DOI:** 10.1101/2024.03.22.24304707

**Authors:** Ahmad Asdo, Alishah Mawji, Isaac Omara, Ivan Aine Aye Ishebukara, Clare Komugisha, Stefanie K. Novakowski, Yashodani Pillay, Matthew O. Wiens, Samuel Akech, Florence Oyella, Abner Tagoola, Niranjan Kissoon, J. Mark Ansermino, Dustin Dunsmuir

## Abstract

**Background:** Pneumonia is the leading cause of death in children globally. In low- and middle-income countries the diagnosis of pneumonia relies heavily on an accurate assessment of respiratory rate, which can be unreliable in nurses and clinicians with less advanced training. In order to inform more accurate measurements, we investigate the repeatability of the RRate app used by nurses in district hospitals in Uganda.

**Methods:** This secondary analysis included 3679 children aged 0-5 years. The dataset had two sequential measurements of respiratory rate using the RRate app. We measured the agreement between respiratory rate observations and clustering around fixed thresholds defined by WHO for fast breathing, which are 60 breaths per minute (bpm) for under two months (Age-1), 50 bpm for two to 12 months (Age-2), and 40 bpm for 12.1 to 60 months (Age-3). We then assessed the repeatability of the paired measurements using the Intraclass Correlation Coefficient (ICC).

**Results:** The respiratory rate measurement took less than 15 seconds for 7,277 (98.9%) of the measurements. Despite respiratory rates clustering around the WHO fast-breathing thresholds, the breathing classification based on the thresholds was changed in only 12.6% of children. The mean (SD) respiratory rate by age group was 60 (13.1) bpm for Age-1, 49 (11.9) bpm for Age-2, and 38 (10.1) for Age-3, and the bias (Limits of Agreements) were 0.3 (-10.8 – 11.3), 0.4 (-8.5 – 9.3), and 0.1 (-6.8, 7.0) for Age-1, Age-2, and Age-3 respectively. Most importantly, the repeatability of the two respiratory rate measurements for the 3,679 children was high, with an ICC value (95% CI) of 0.95 (0.94 – 0.95).

**Discussion:** The RRate measurements were both efficient and repeatable. The simplicity, repeatability, and efficiency of the RRate app used by healthcare workers in LMICs supports more widespread adoption for clinical use.

## Introduction

Pneumonia is the leading cause of death in children worldwide, claiming the lives of 725,000 children under the age of 5 every year, including 190,000 neonates.^1,2^ Sub-Saharan Africa accounts for 30% of this global burden.^3^ Many of these pneumonia deaths are preventable with accurate diagnosis and prompt treatment.

The World Health Organization (WHO) Integrated Management of Childhood Illness (IMCI) guidelines rely significantly on clinical respiratory rate measurement for diagnosing and managing pneumonia in Low- and Middle-Income Countries (LMIC). However, respiratory rate remains difficult to measure accurately despite its profound clinical importance.^4^ First level sick-child support in LMICs is primarily done by nurses with no routine access to more sophisticated diagnostic tools.^5^ Nurses in these settings have been shown to make less-sensitive identifications of pneumonia compared to clinicians because they do not receive tailored training on pneumonia detection.^6^

The “true” underlying physiological respiratory rate is ephemeral and time-varying; however, one factor that can be measured and optimized in clinical practice is repeatability. Repeatability is the consistency between different sets of measurements taken under similar conditions.^9^ The uncertainty in respiratory rate measurement introduced by poor repeatability will significantly reduce the reliability of clinical decisions.^10^ However, a simple, efficient, and repeatable method of measuring respiratory rate is likely to improve the accuracy of respiratory rate measurements by nurses in LMICs.

The WHO recommends the Acute Respiratory Infection (ARI) timer for measuring respiratory rate.^7^ Unfortunately, the usability of the ARI is suboptimal when caring for a restless child who may be moving, crying, or breathing rapidly. ^8^ In addition, it takes a minimum of a full minute to measure RR with the ARI timer, which is difficult in a busy understaffed clinic or emergency department.^8^ RRate is a smartphone application for measuring respiratory rate in LMIC hospital outpatient departments. Previous research has optimized the trade-off between usability and accuracy and compared the RRate app to the ARI timer in a controlled setting.^11,12^ The objective of this study was to evaluate the repeatability of RRate when used to measure respiratory rate in a busy outpatient department of a Ugandan hospital.

## Methods

Data for this secondary analysis was collected during the baseline phase of a multisite implementation study for a digital triaging platform, Smart Triage (Clinical Trials.gov Identifier: NCT04304235).^13,14^ Measurements were obtained from 4,604 children who presented to the outpatient department of Jinja Regional Referral Hospital (JRRH, 1,748 children) and Gulu Regional Referral Hospital (GRRH, 2,856 children) in Uganda from April, 27^th^ 2020, to April 16^th^ 2022. JRRH and GRRH have admission rates of 20% and 18% respectively and both are public regional referral hospitals in an LMIC. Measurements were performed by nurses trained in the use of the RRate application.

## Data collection

After consent, a nurse collected over 200 variables, including clinical signs, symptoms, and sociodemographic variables.^13^ For each child, two measurements of respiratory rate were taken using the RRate app. The user observes the patient’s chest and taps the touch screen on the onset of each inhalation. Inter-breath intervals are then calculated from the time between taps. The RRate app ensures the user taps consistently five times.^15^

## Data Analysis

This secondary analysis only included children up to 5 years of age. We excluded children without both respiratory rate measurements, those whose paired respiratory rate measurements that were more than five minutes apart, or those who had more than 80% of their clinical data missing. We used Bland Altman plots to assess systematic errors as well as bias and limits of agreement between the first respiratory rate measurement (RR-1) and the second respiratory rate measurement (RR-2). We investigated the respiratory rates clustering around the thresholds for fast breathing, suggested by the WHO.^4^ The thresholds for fast breathing are 60 bpm for children younger than two months (Age-1), 50 bpm for children between two to 12 months (Age-2) and 40 bpm for those between 12.1 to 60 months (Age-3). Additionally, admission rates were compared between age groups using Analysis of Variation (ANOVA).

The Intraclass Correlation Coefficient (ICC) was assessed as a measure of repeatability between RR-1 and RR-2. ICC values have been defined as follows: less than 0.2—slight repeatability, between 0.2 and 0.4—low repeatability, between 0.4 and 0.7—moderate repeatability, between 0.7 and 0.9—high repeatability, and greater than 0.9—very high repeatability. ^16^ We first calculated an ICC for each observer (healthcare worker) using a single rater model. We then calculated the ICC using all measurement pairs with a multiple-rater model to account for any probabilistic dependency between the observers.

## Ethical considerations

The parent study was approved by the institutional review boards at the University of British Columbia in Canada (ID: H19-02398; H20-00484), the Makerere University School of Public Health in Uganda (ID: 743) and the Uganda National Council for Science and Technology (ID: HS528ES). All participants consented to use of their data for secondary analyses. A parent or guardian provided written informed consent prior to enrollment. In addition, assent was required from children aged older than 8 years. Data for the retrospective study was accessed on January 27, 2022, and there was no access to identifying patient data.

## Results

We reviewed 4,604 children from the two facilities and excluded 925 who did not meet the eligibility criteria. A total of 3,679 paired observations were analyzed (Figure 1). The median (IQR) time difference between the two respiratory rate measurements was 57 (25 – 91) seconds.

**Figure 1.**
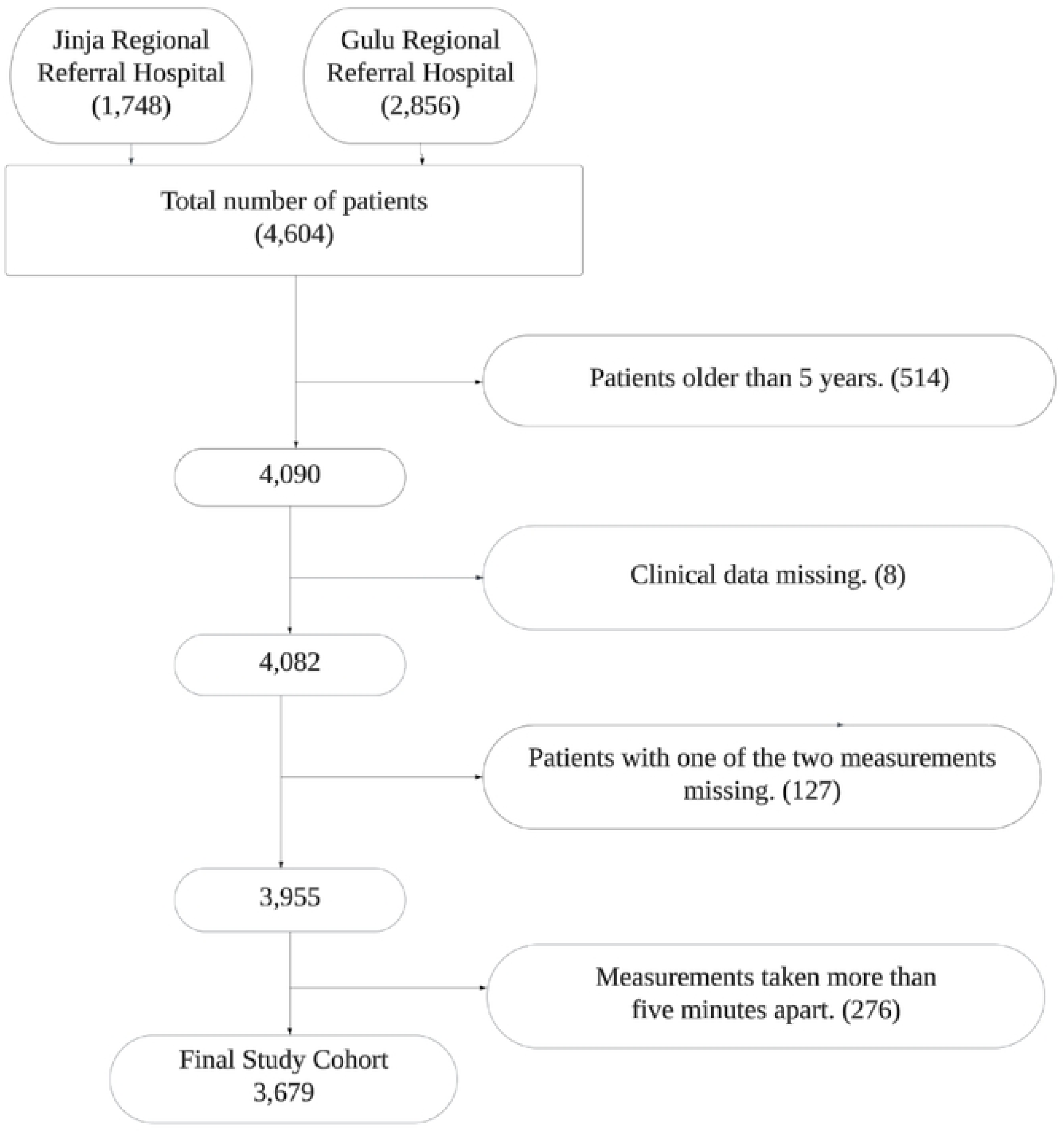
Consort diagram for children included in the study.

The mean (SD) respiratory rate was 44 (12.9) bpm for all children. The mean respiratory rate by age group was within two breaths of the corresponding WHO threshold for fast breathing (Table 1). The admission rate was lower for Age-2 children (Table 1 and Appendix).

**Table 1.**
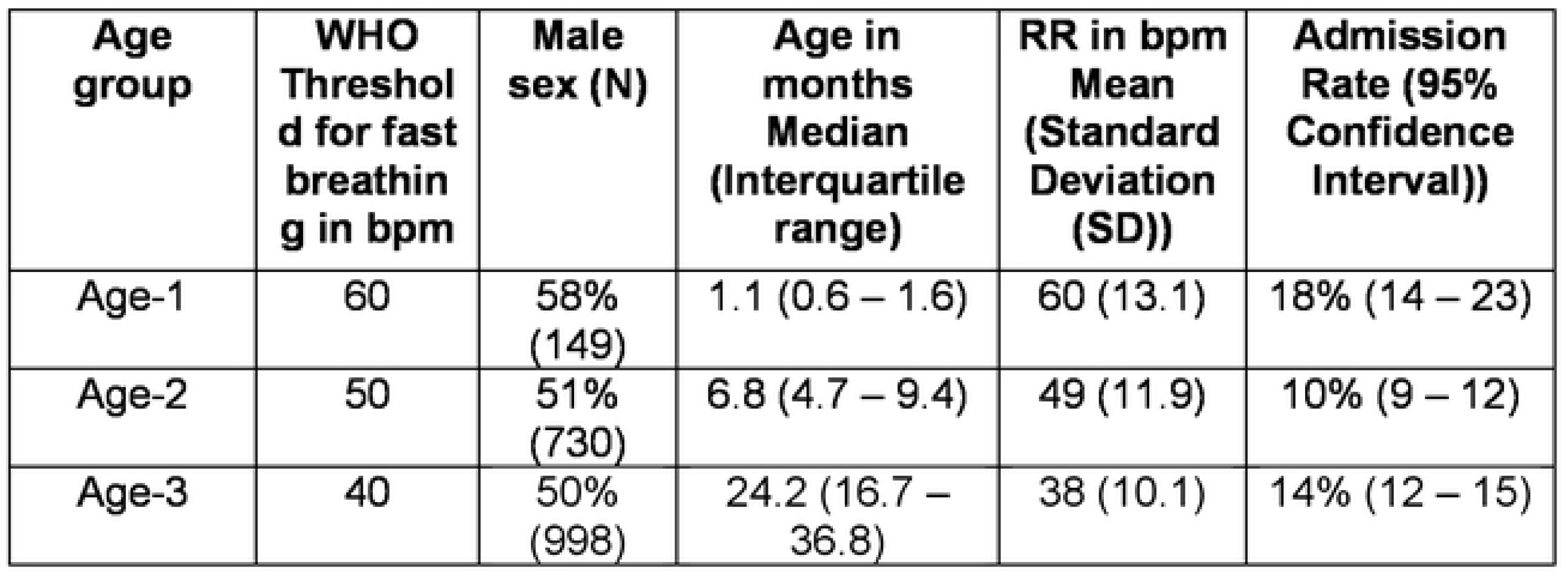
Demographics and parameters of respiratory rate for the study cohort.

| **Age group** | **WHO Threshold for fast breathing in bpm** | **Male sex (N)** | **Age in months Median (Interquartile range)** | **RR in bpm Mean (Standard Deviation (SD))** | **Admission Rate (95% Confidence Interval))** |
| --- | --- | --- | --- | --- | --- |
| Age-1 | 60 | 58% (149) | 1.1 (0.6 – 1.6) | 60 (13.1) | 18% (14 – 23) |
| Age-2 | 50 | 51% (730) | 6.8 (4.7 – 9.4) | 49 (11.9) | 10% (9 – 12) |
| Age-3 | 40 | 50% (998) | 24.2 (16.7 – 36.8) | 38 (10.1) | 14% (12 – 15) |

Study nurses were able to measure respiratory rates from the first 5 breaths (5 taps on the screen, 4 inter-breath intervals) in 6,189 (84.1%) of measurements. For 6,909 (93.9%) measurements, the nurses required 6 or less breaths (6 or less taps) for measurement. A respiratory rate measurement was obtained in less than 15 seconds in 7,277 (98.9%) of the total measurements completed.

A Bland Altman plot for all the children showed strong agreement between RR-1 and RR-2 with a bias of 0.24 breaths per minute (Figure 2). The limits of agreements were the widest in Age-1 and narrowed with increasing patient age. Additionally, bias was lowest in Age-3. (Figures 3-5). There were 463 children (12.6%) who had their respiratory rate classification changed according to IMCI (Table 2).

**Figure 2.**
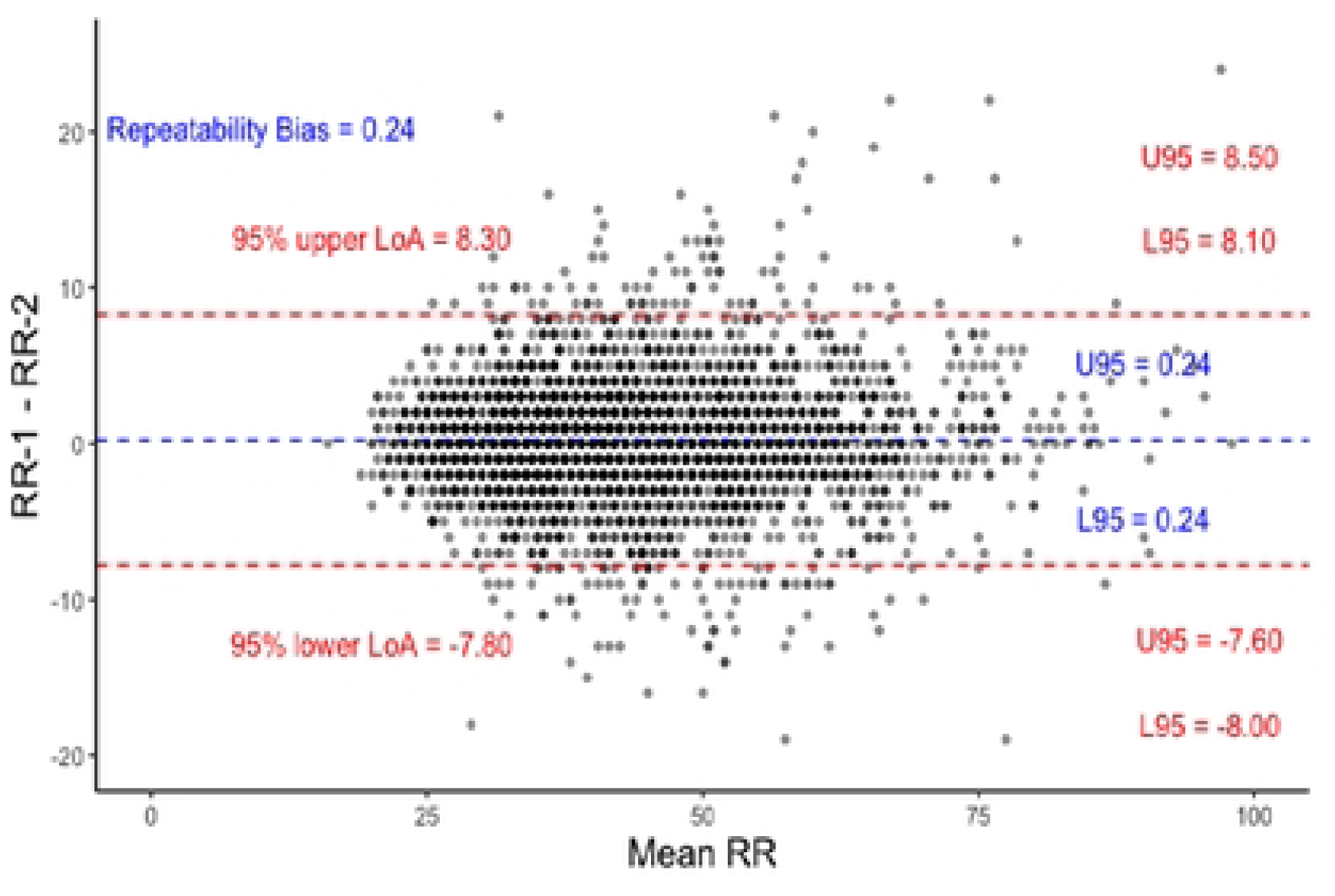
Bland Altman plot for RR-1 and RR-2 across all children. Limits of Agreement (LoAs) and bias are under repeatability conditions, considering one set of samples as the test and the other as the reference.

**Figure 3.**
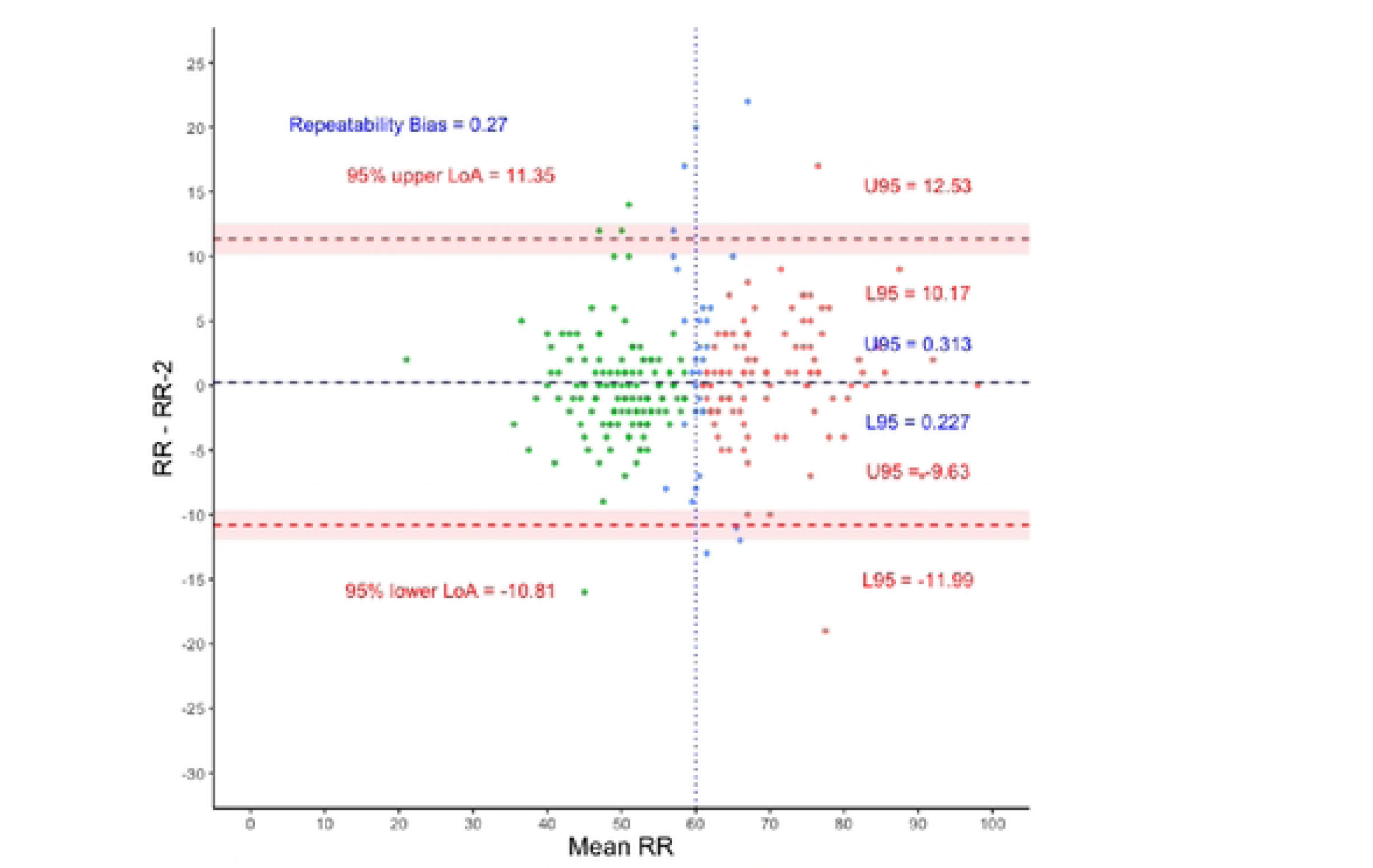
Bland Altman plot for Age-1. N = 259. Threshold for fast breathing is 60 bpm. Red dots represent children with two fast breathing measurements, green dots - two normal breathing, and blue dots - one fast and one normal.

**Figure 4.**
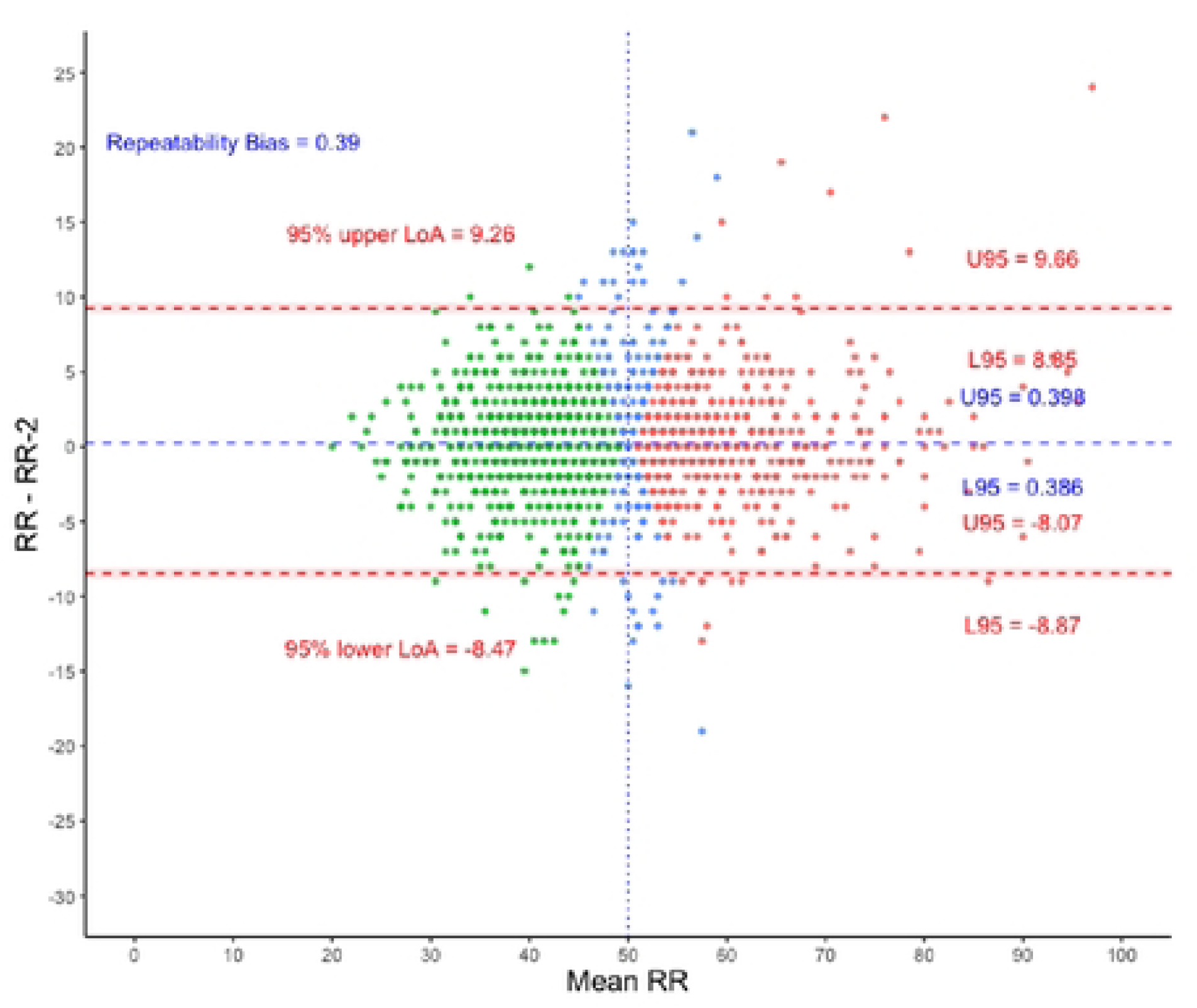
Bland Altman Plot for Age-2. N = 1,428. Threshold for fast breathing is 50 bpm. Red dots represent children with two fast breathing measurements, green dots - two normal breathing, and blue dots – one fast and one normal.

**Figure 5.**
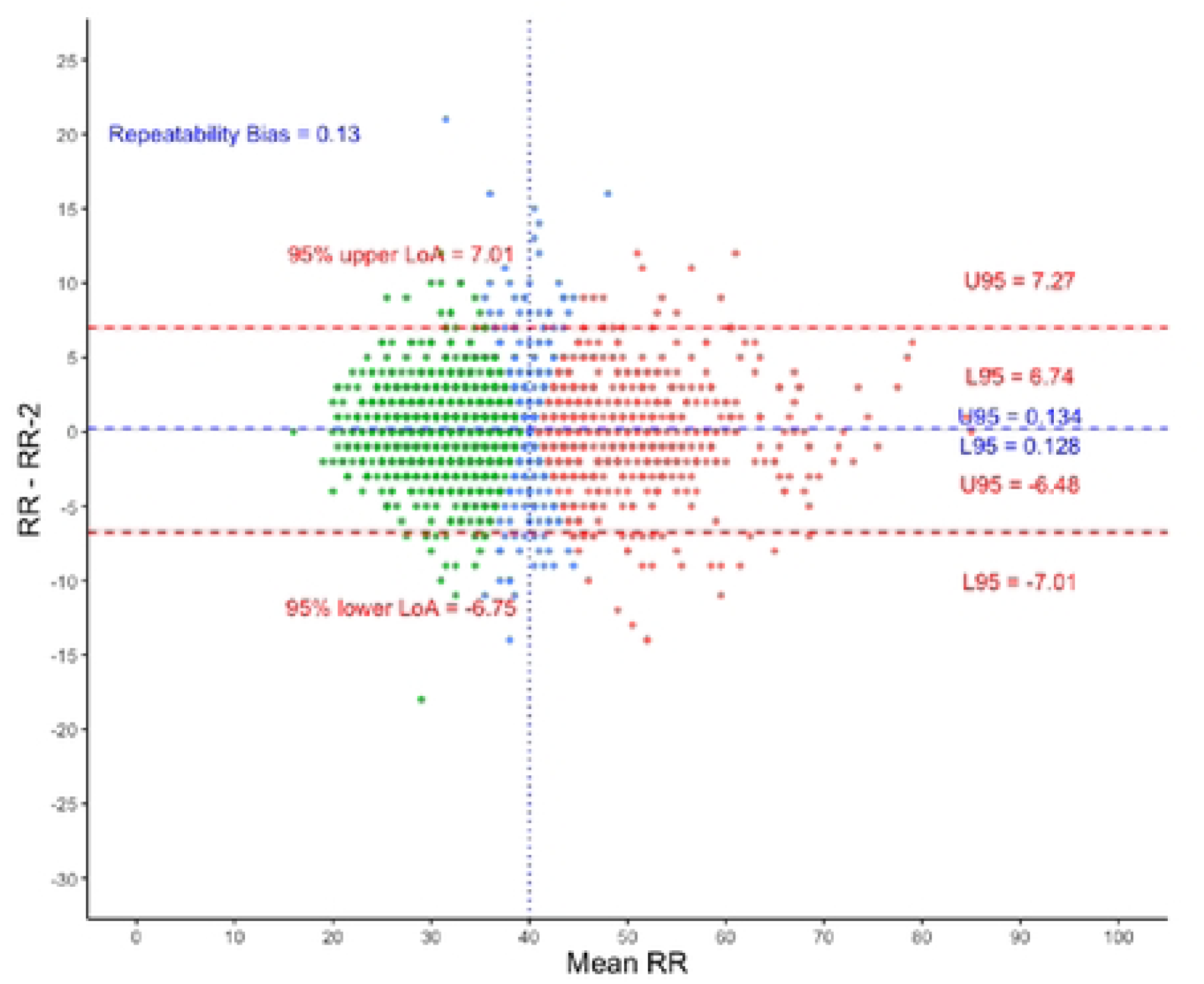
Blond Altman Plot for Age-3. N = 1,992. Threshold for fast breathing is 40 bpm. Red dots represent children with two fast breathing measurements, green dots - two normal breathing, and blue dots – one fast and one normal.

**Table 2.** Children’s breathing classification based on age group and WHO thresholds.

|  | Percentage of Age-1 | Percentage of Age-2 | Percentage of Age-3 | All children |
| --- | --- | --- | --- | --- |
| Two Normal Measurements | 46.7% | 51.3% | 57.0% | 54% |
| Two Fast Measurements | 40.5% | 35.4% | 30.9% | 33.4% |
| One Normal | 12.7% | 13.3% | 12.0% | 12.6% |

The repeatability performance between RR-1 and RR-2, indicated by the ICC, was ≥ 0.90 for 12 of 14 users despite the variable sample size measured by each user (Table 3). The overall ICC (IQR) of the two respiratory rate measurements was 0.95 (0.94 – 0.95). This indicates the very high repeatability of the RRate app.

**Table 3.**
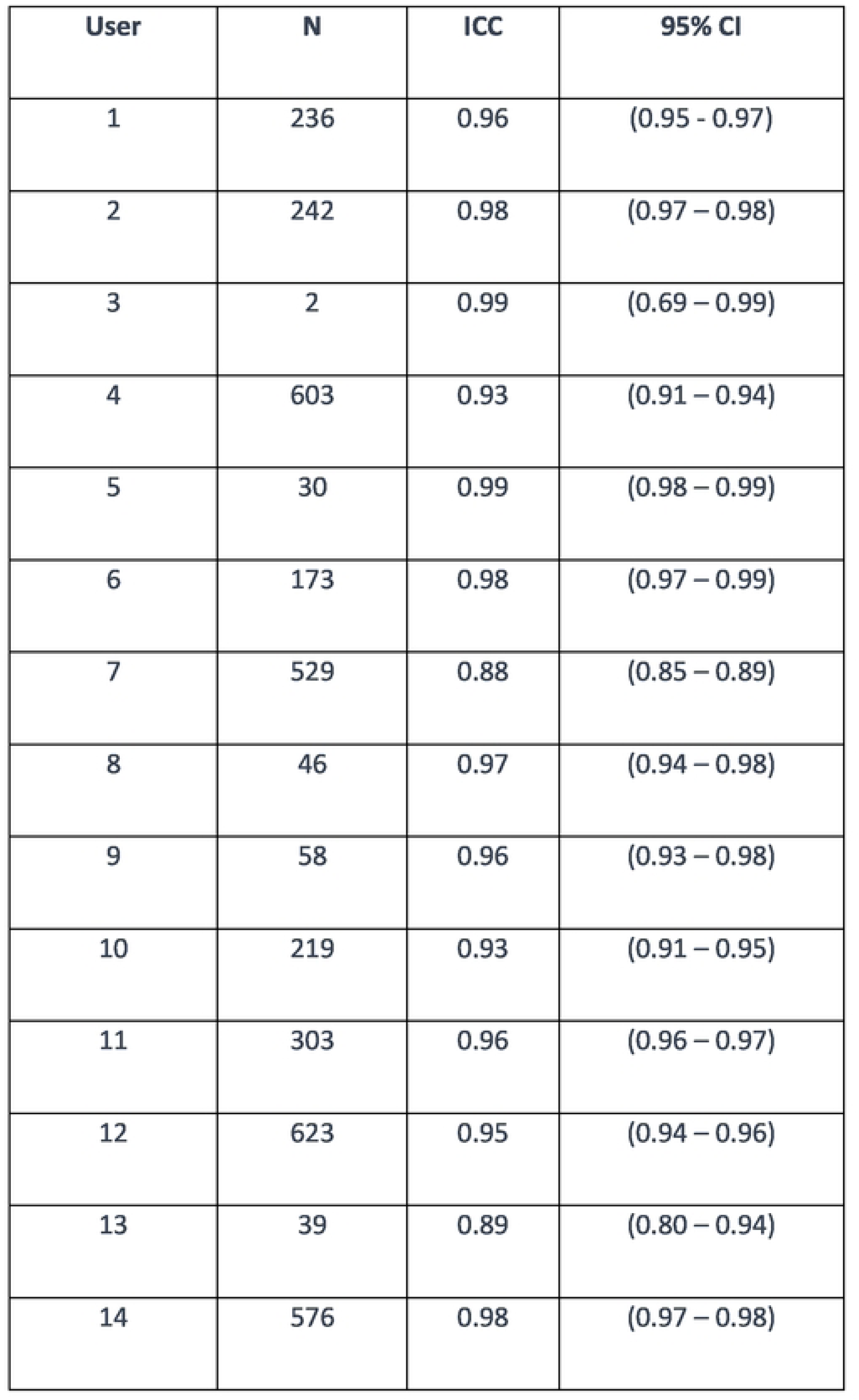
Intraclass Correlation Coefficient /ICC) per user.

| User | N | ICC | 95% CI |
| --- | --- | --- | --- |
| 1 | 236 | 0.96 | (0.95 - 0.97) |
| 2 | 242 | 0.98 | (0.97 – 0.98) |
| 3 | 2 | 0.99 | (0.69 – 0.99) |
| 4 | 603 | 0.93 | (0.91 – 0.94) |
| 5 | 30 | 0.99 | (0.98 – 0.99) |
| 6 | 173 | 0.98 | (0.97 – 0.99) |
| 7 | 529 | 0.88 | (0.85 – 0.89) |
| 8 | 46 | 0.97 | (0.94 – 0.98) |
| 9 | 58 | 0.96 | (0.93 – 0.98) |
| 10 | 219 | 0.93 | (0.91 – 0.95) |
| 11 | 303 | 0.96 | (0.96 – 0.97) |
| 12 | 623 | 0.95 | (0.94 – 0.96) |
| 13 | 39 | 0.89 | (0.80 – 0.94) |
| 14 | 576 | 0.98 | (0.97 – 0.98) |

## Discussion

In this secondary analysis we found a high repeatability of respiratory rate measurements taken by nurses using the RRate application on children arriving to two busy public referral hospitals in Uganda. The Bland Altman plot (Figure 2) further indicates the strong agreement between the two measurements with a bias of only 0.24 bpm. Additionally, the app measurements were completed rapidly, as 98.9% of the measurements took less than 15 seconds.

The RRate app is more efficient in LMICs settings compared to other devices. The RRate app and the ARI timer have previously been shown to have similar accuracy in a controlled setting where de-identified videos of anesthetized children were used as a standard.^12^ However, RRate provides a respiratory rate in children in 15 seconds versus 60 seconds required using the ARI timer; a distinct advantage in settings where healthcare workers need to efficiently triage large numbers of children rapidly.^17^ During the measurement, RRate does not require the child to lie motionless. Another attractive feature of the RR app is its interoperability with an array of mobile devices that can run this application at no additional cost.

A previous study has shown a lack of accuracy in the RRate app when using a video-based counting method as a reference. However, there was no formal training in the use of RRate and repeatability was not reported.^19^ Those findings were in contrast to another analysis, where RRate showed excellent agreement with the video-based counting reference.^11^

Evaluating respiratory rate measurement in experimental controlled settings may not be comparable to real-world settings. However, the demonstration of robust repeatability would be highly desirable for real world applications. Automated respiratory rate devices such as Masimo Rad G pulse oximeter have also been evaluated in real-world settings and have been shown to have variable accuracy with wide 95% LoAs (-34 – 6). However, automated device repeatability has not been demonstrated.^18^

## Implications for Clinical Care

Despite the importance of respiratory rate measurement, measuring an accurate rate is an ongoing challenge to healthcare workers.^4^ Respiratory rate assessment is relied upon to make clinical decisions in LMICs where the burden of pneumonia is highest and where nurses are heavily relied upon to identify pneumonia. Considering the relatively lower sensitivity of decisions made by nurses compared to those made by clinicians, utilizing the RRate mobile app can enhance the capabilities of nurses who heavily depend on respiratory rate to identify rapid breathing.^6^ It will also inform decisions to implement pneumonia treatment protocols. The low cost and accessibility of RRate makes it appropriate for use in low-resource settings, especially when there are many children and few healthcare workers.

## Limitations

The generalizability of these results may be limited since the data is from only two institutions and only includes 14 observers. The time needed to measure RRate is reported starting from when the first breath is observed (tapped). This does not include the time taken to prepare for the measurement, which may include revealing the child’s chest or calming them. However, using a traditional method of counting breaths would also not start until after a child was calmed. In situations where the user failed to tap the screen consistently for 12 breaths, the app would prompt the user to redo the measurement and would not record the time taken for the failed measurement. However, given that 84.1% of measurements needed only the initial five breaths, and 93.9% needed a maximum of six breaths, it is likely that very few (if any) children needed more than one attempt.

## Conclusion

The RRate app is an open-source and free solution to respiratory rate measurement with very high repeatability and agreement between measurements. RRate is an efficient and repeatable alternative to the breath counting method. The screen tapping method could be incorporated into medical monitoring devices to assess respiratory rate.

## Data Availability

Data is available upon a reasonable request from the corresponding author.

https://doi.org/10.5683/SP3/XO7BVV

## Acknowledgements

We express our gratitude to the Smart Triage research staff at Jinja and Gulu Regional Referral Hospitals for their assistance in data collection and staff at WALIMU for assisting with study setup and equipment procurement. We also extend our appreciation to all the participating children and caregivers in the study. Special thanks to Nurse Collins Agaba for her valuable contribution in project coordination.

